# Antibodies anti-SARS-CoV2 time-course in patients and vaccinated subjects: an evaluation of the harmonization of two different methods

**DOI:** 10.1101/2021.08.28.21262543

**Authors:** Ruggero Dittadi, Mara Seguso, Isabella Bertoli, Haleh Afshar, Paolo Carraro

## Abstract

The time-course of antibodies anti SARS-CoV2 is not yet well elucidated, especially in people who underwent a vaccination campaign. In this study we measured antibodies anti-S1 and anti-RBD with two different methods both in patients and in vaccinated subjects.

108 specimens from 48 patients diagnosed as COVID-19 affected (time from the onset of symptoms from 3 to 368 days) and 60 specimens from 20 vaccinated subjects (collected after 14 days from the first dose, 14 days and 3 months after a second dose of Comirnaty) were evaluated.

We used an ELISA method that measure IgG against anti-Spike 1 and a chemiluminescence immunoassays that measure IgG anti-RBD.

In the patients, antibodies concentrations tend to decline after a few months with both methods, but persist relatively high up to nearly a year after symptoms.

In vaccinated subjects, antibodies were already detectable after the first dose, but after the booster they show a significant increase. However, the decrease is rapid, given that after 3 months after the second vaccination they are reduced to less than a quarter.

The conversion of the results into BAU units improves the relationship between the two methods. However, in vaccinated subjects there was no evidence of proportional error after the conversion, while in the patients the difference between the two methods remained significant.

## 1. Introduction

The determination of the antibodies against SARS-CoV-2 could be useful in epidemiological studies for estimating the spread of the infection and the lethality rate, in the serological diagnosis for individuals with mild or moderate symptoms and asymptomatic, in the first screening of convalescent patients for plasma collection and in monitoring of the antibody response of vaccinated subjects.

Long term time-course of antibodies response in Covid-19 disease is not yet fully determined. Some studies show a significant decrease of antibodies concentrations within 3-4 months from the onset of symptoms [1-4]. Others reports find constant or only slightly decreased levels starting from 4 months and up to 10 months from the symptoms’ onset [5-8], even when specific neutralizing antibodies [9] were measured. In particular, the time-course of the antibody response seems variable also according to the method used [8,10]. On the other hand, antibodies seem to persist through 4-6 months in vaccinated subjects [11,12].

The aim of this work is to evaluate the performance of 2 methods for the determination of antibodies to SARS-CoV-2 in patients with a long-term time-course. We also evaluated the presence of antibodies in a little cohort of subjects vaccinated with Comirnaty vaccine.

## 2. Materials and Methods

We recruited symptomatic subjects who presented at the dell’Angelo Hospital (Mestre, Italy) in March 2020, resulted affected by COVID-19 according to both clinical and laboratory criteria. Forty-eight patients with known date of symptoms’ onset were included in the study (41 males, 7 females, median age 62.3 years, minimum 28, maximum 87). The median time from the onset of symptoms was 26 days (minimum 3, maximum 368). A total of 108 withdrawals were collected. Number of withdrawals per patient and other patients’ characteristics were reported in table 1. Moreover, serum samples were collected from 20 healthcare workers after 14 days from the first dose, 14 days and 3 months after a second dose of Comirnaty vaccine (BNT162b2, BioNTech/Pfizer, Mainz, Germany/New York, United States). All subjects underwent periodical nasopharyngeal swab testing (every 2 or 3 weeks) and resulted negative to the antibodies determination prior to vaccine administration.

**Table 1.** Characteristics of the studied patients. The disease severity was classified according the WHO guidance “Laboratory testing for coronavirus disease (COVID-19) in suspected human cases”)

| Symptoms at the onset of disease | Frequency (%) |
| --- | --- |
| Fever | 75.0 |
| Cough | 60.4 |
| Dyspnoea | 29.2 |
| Nausea | 8.3 |
| Asthenia | 6.3 |
| Others | 10.5 |
| Disease severity | n of patients |
| Mild | 9 |
| Moderate | 16 |
| Severe | 11 |
| Critical | 12 |
| n of withdrawals | n patients |
| 1 | 19 |
| 2 | 12 |
| 3 | 7 |
| 4 | 7 |
| 5 | 2 |
| 6 | 1 |

### The specimens were stored at -80°C until the assay

IgG were measured with an ELISA method, the anti-SARS-CoV-2 QuantiVac ELISA IgG (Euroimmun, Lubeck, Germany) and a two-step chemiluminescence microparticle immunoassays SARS-CoV-2 IgG anti-RBD (SNIBE, Shenzen, China).

Both the assays were performed according to the manufacturer’s instructions.

The good analytical characteristics of the two methods and the satisfactory correlation with the neutralization tests were previously evaluated and confirmed [13-15]. The concentrations were measured taking into consideration the previously determined linearity of the respective methods [16].

In the ELISA Euroimmun method, the concentrations of the antibodies against the S1 protein were determined through the interpolation with a six-point calibration curve (from 1 to 120 Relative Units/mL). Results <8 RU/mL were considered as negative, results >8 RU/mL and ≤11 RU/mL were considered as indeterminate and >11 RU/mL as positive.

The IgG anti-RBD method measure antibodies against receptor binding domain of the S1 protein, were carried out on the analyser Maglumi 800 (SNIBE, Shenzen, China), and use a nine-point master curve (from 1 to 100 Arbitrary Units/mL) periodically adjusted by a 2-point calibration. Results >= 1 AU/mL were considered as positive.

The statistical analyses were performed with MedCalc © Software, Version 7.4.2.0 (MedCalc Software, Mariakerke, Belgium).

## 3. Results

The overall concordance rate between methods was 89.8% (kappa statistics, 0.54; 95% CI, 0.30–0.78). The results of patients’ specimens were subdivided into 7 groups, for which the qualitative performance was evaluated (Table 2).

**Table 2.**
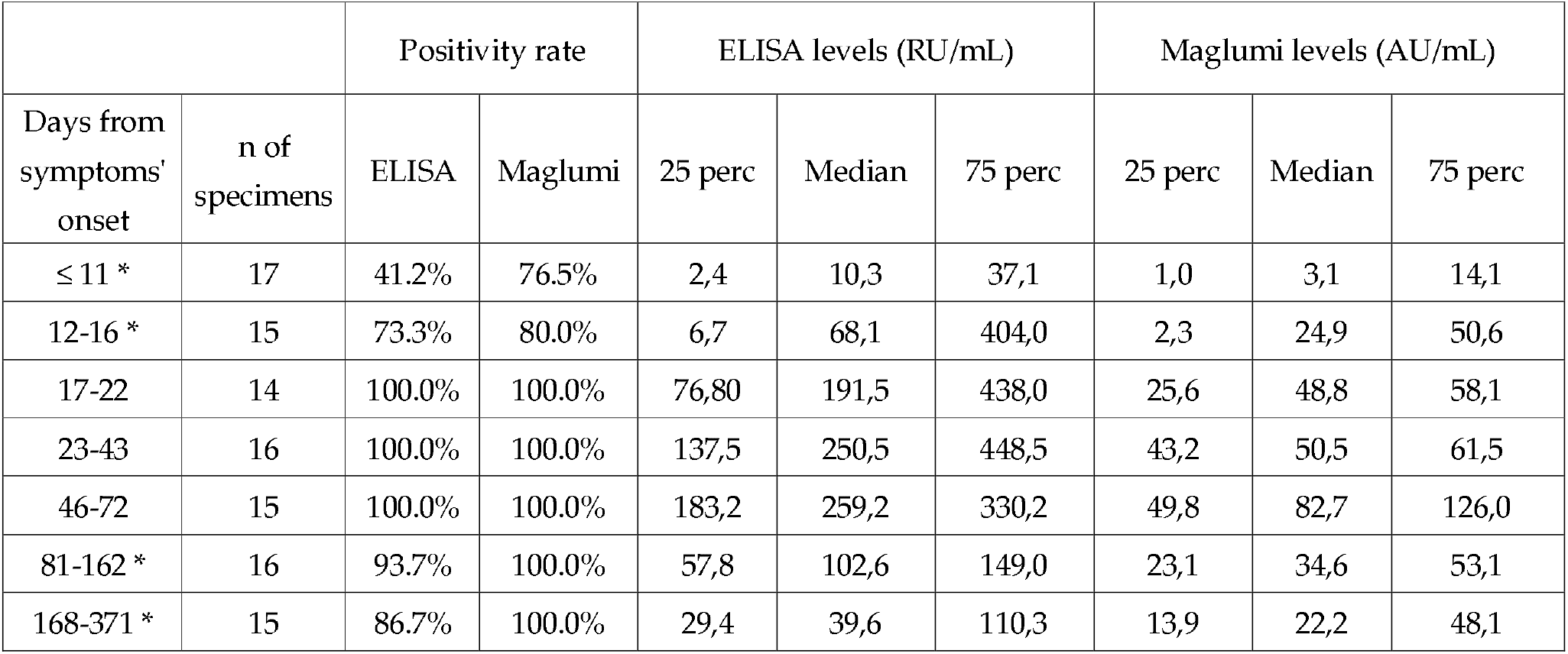
Sensitivity and antibodies levels of ELISA and Maglumi methods in the different patient’s specimens subdivided in time frames according to the day from the onset of symptoms Asterisks represent the classes of cases significantly different from that with higher concentrations.

The quantitative relationship showed a satisfactory correlation, although with relative disperse distribution of cases. Passing-Bablock regression resulted “*Maglumi= 0*.*284 (0*.*24/0*.*33)* +*0*.*581 (−0*.*21/2*.*64) ELISA*” (Fig. 1 A).

**Figure 1.**
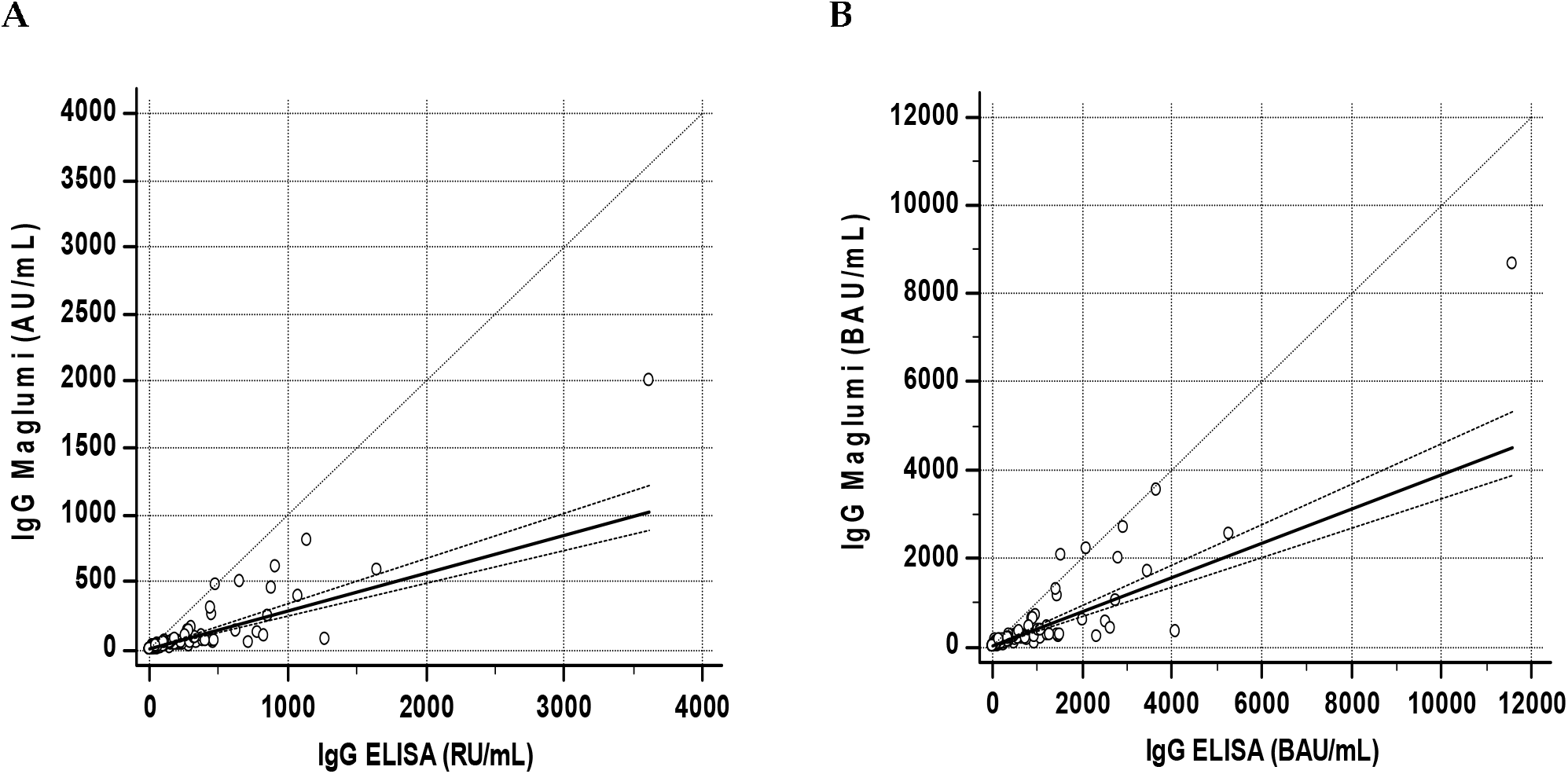
Correlations between ELISA and Maglumi SARS-CoV-2 IgG levels in patients’ specimens. The trend lines represent the Passing-Bablock correlation **A** Concentrations expressed in Arbitrary Units [*Maglumi= 0*.*581 (−0*.*21/2*.*64) ELISA* + *0*.*284 (0*.*24/0*.*33)*] **B** Concentrations expressed as BAU [*Maglumi = 2*.*45 (−1*.*6* / + *10*.*6)* +*0*.*39 (0*.*33 / 0*.*46) ELISA]*

The differences in concentration between age groups were statistically significant for both methods (Kruskall-Wallis test p=0.00002 for both Maglumi and ELISA).

The antibodies’ levels showed a similar time-course with the two methods. After a rapid increase, the concentrations begin to decrease slightly after about 80-100 days (figure 2 and 3).

**Figure 2.**
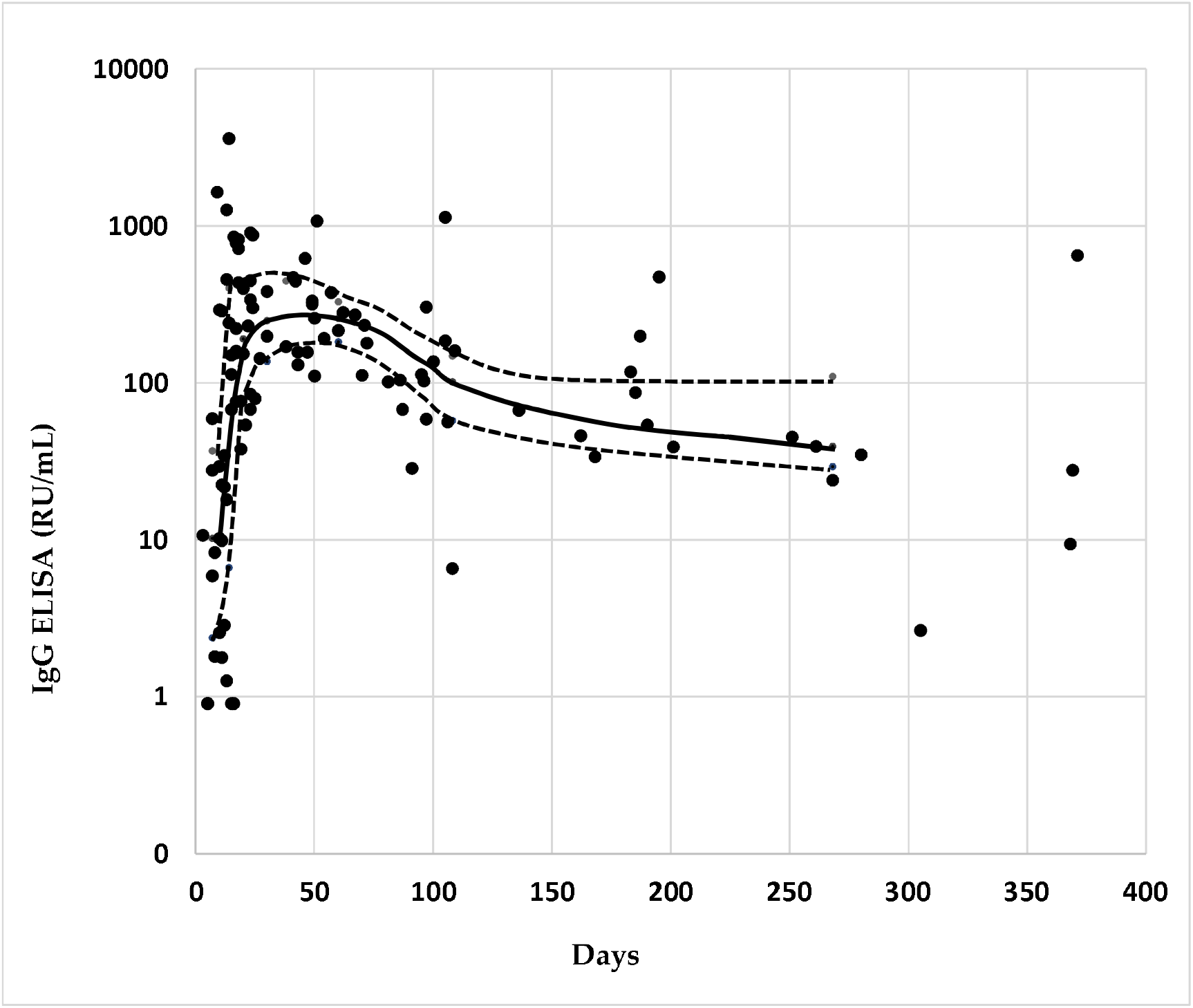
Distribution of IgG levels of the single specimens measured by ELISA in relation to the days since the onset of symptoms. In abscissa are reported the days from the onset of symptoms, in ordinate the concentrations of IgG. The solid line connects the median concentrations of IgG for each class of cases, the dotted line connects the respective 25°-75° percentile.

**Figure 3.**
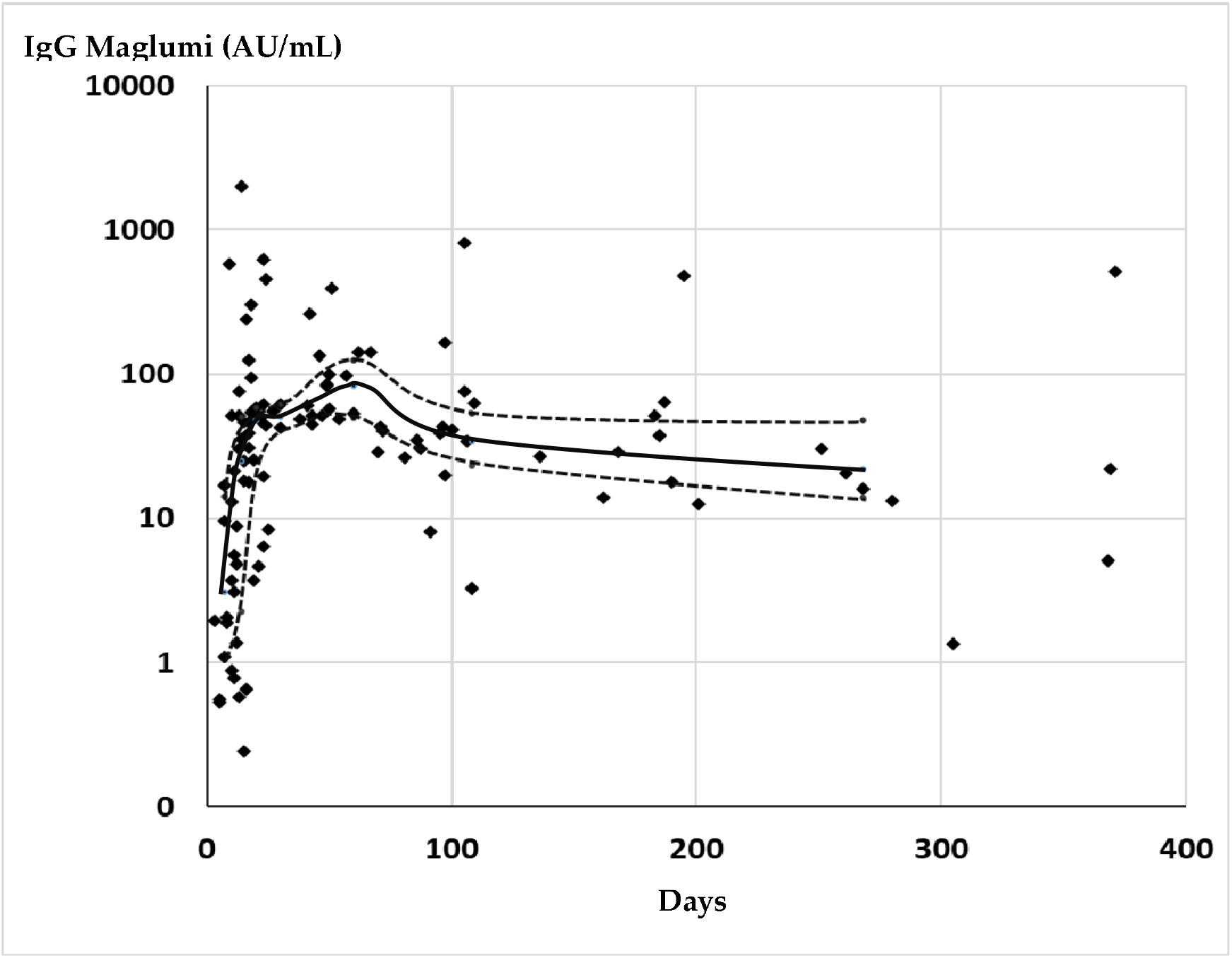
Distribution of IgG levels of the single specimens measured by Maglumi in relation to the days since the onset of symptoms. In abscissa are reported the days from the onset of symptoms, in ordinate the concentrations of IgG. The solid line connects the median concentrations of IgG for each class of cases, the dotted line connects the respective 25°-75° percentile.

The ELISA method showed a sensitivity of about 87% after 180 days, while the sensitivity of Maglumi remains 100%.

The results of the determination with the two methods in the 13 patients with more than one blood collection at least up to 180 days after the onset of symptoms were shown in figure 4.

**Figure 4.**
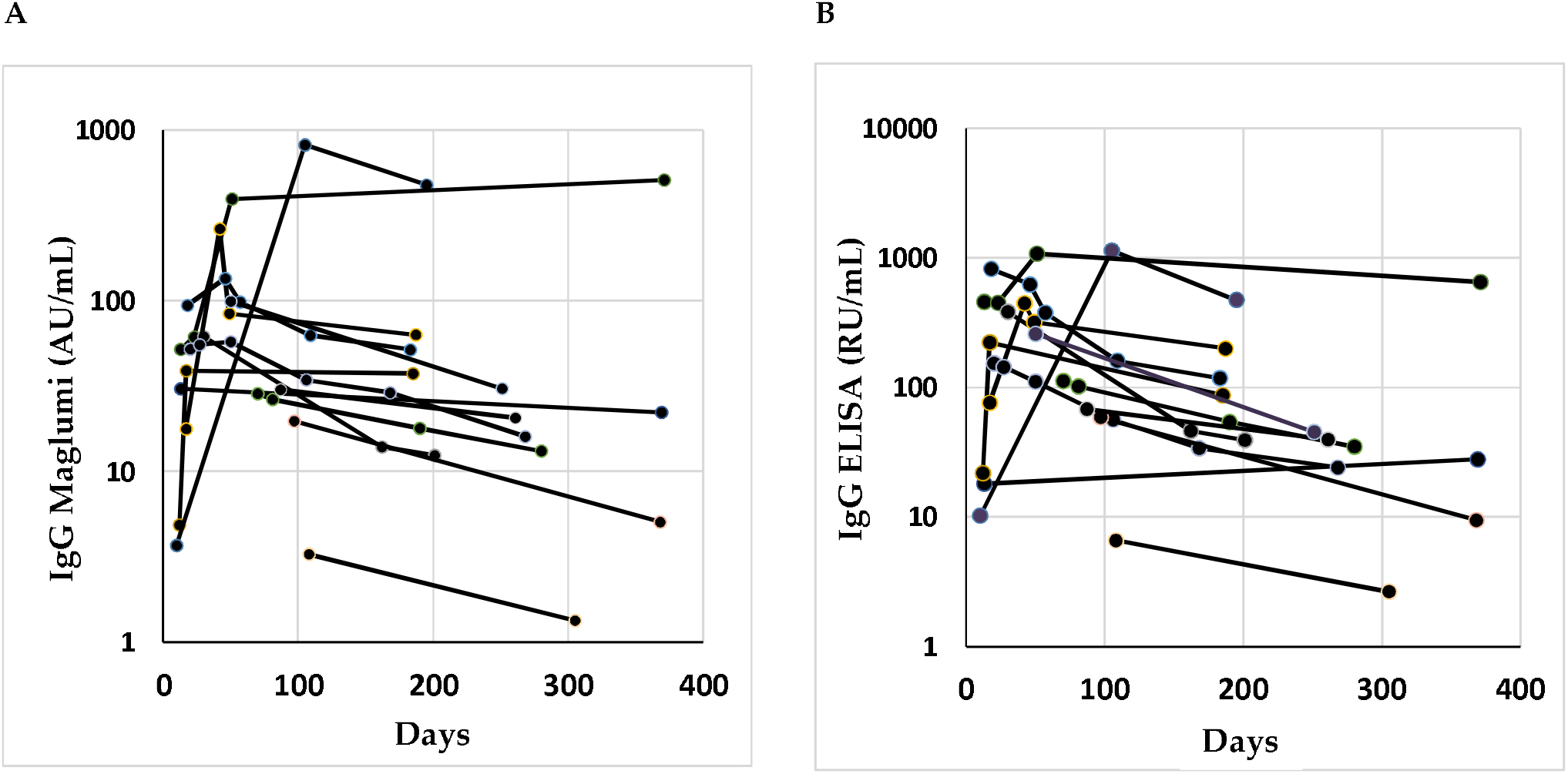
Spaghetti plot of the 13 patients with more than one withdrawal in more than 180 days from the onset of symptoms, measured by Maglumi (**A**) and ELISA (**B**).

All the vaccinated subjects were positive 15 days after the first inoculum with Maglumi method, while with the ELISA method 4/22 cases resulted negative.

Fifteen days after the booster all the samples were positive with both methods, and the concentrations resulted more than 20 times the first withdrawal (table 3).

**Tab. 3.** Concentrations of antibodies anti SARS-CoV2 in the vaccinated subjects, expressed both in Arbitrary Units and in BAU

|  |  | ELISA levels |  | Maglumi levels |  |
| --- | --- | --- | --- | --- | --- |
|  |  | Median | Interquartile range | Median | 75 perc |
| Arbitrary Units/mL | 15 days after the first dose | 27.2 | 16.4 – 42 | 18.1 | 6.6 - 27.7 |
|  | 15 days after the second dose | 704.5 | 388.5 – 940 | 499 | 335.9 – 756.8 |
|  | 3 months after the second dose | 129.6 | 95.2 – 244 | 84.6 | 47.7 – 195 |
| Binding Antibody Units/mL (WHO) | 15 days after the first dose | 87 | 58.1 – 113.8 | 78.4 | 30.8 – 113 |
|  | 15 days after the second dose | 2254 | 1243 – 3008 | 2160 | 1454 – 3277 |
|  | 3 months after the second dose | 366.1 | 206.7 - 844 | 414.7 | 305 - 781 |

The correlations between the two methods resulted satisfactory, especially after the second dose (figure 5 and 6). The Passing-Bablock regression were: *Maglumi= -0*.*89(−6*.*1*/+*1*.*2)* +*0*.*59 (0*.*47*/*0*.*78) ELISA* for the specimens after the first dose and *Maglumi= - 52*.*4(−107*/+*19*.*2)* +*0*.*85 (0*.*74*/*0*.*92) ELISA* for the specimens after the second administration.

**Figure 5.**
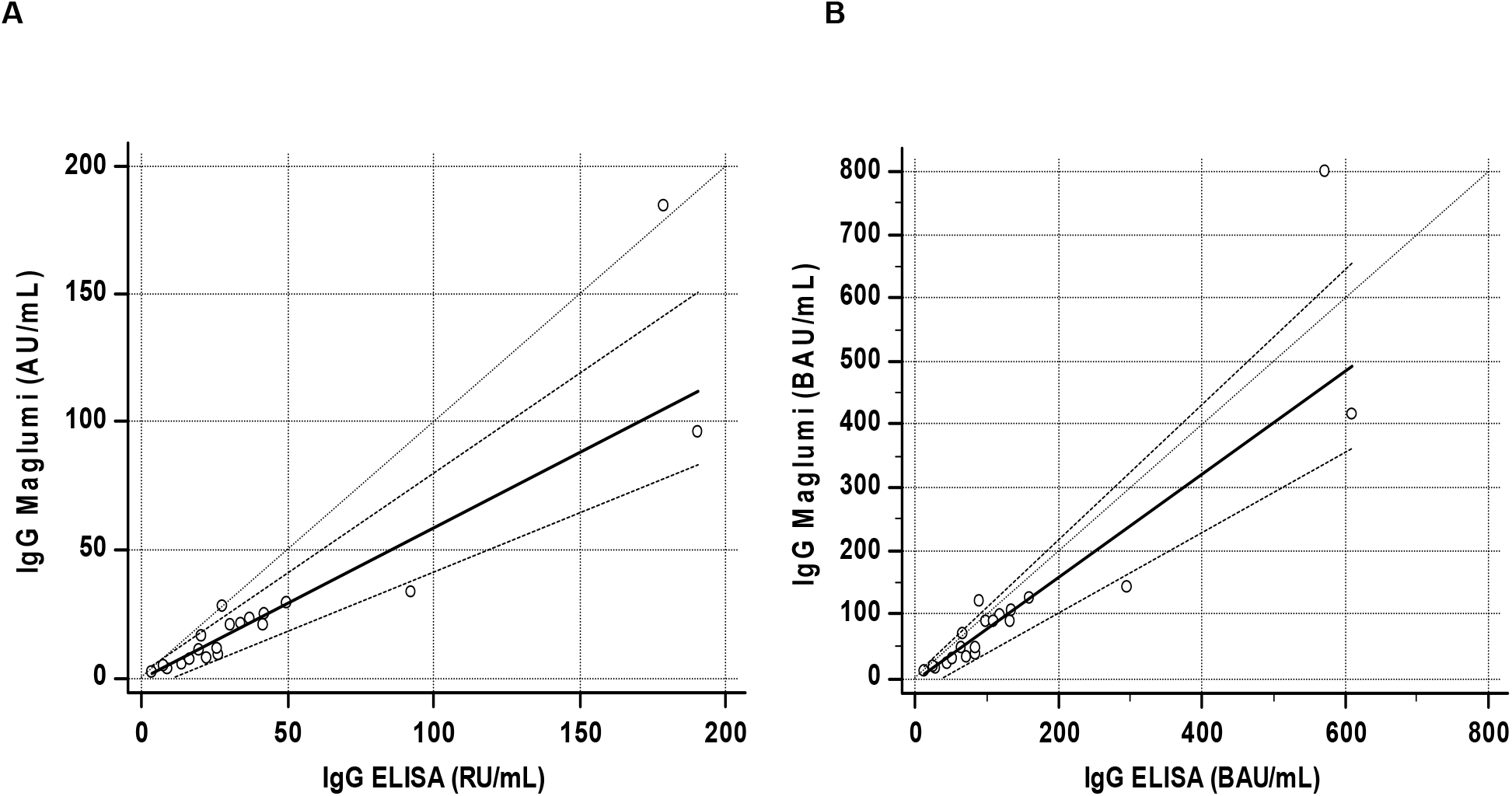
Correlation between ELISA and Maglumi methods in vaccinated subjects after the first dose. The trend lines represent the Passing-Bablock correlation **A** Concentrations expressed in Arbitrary Units [*Maglumi*= *-0*.*89(−6*.*1*/+*1*.*2)* +*0*.*59 (0*.*47*/*0*.*78) ELISA* **B** Concentrations expressed as BAU [*Maglumi* = *-5*.*1 (−26*.*8* / + *5*.*0)* +*0*.*82 (0*.*64* / *1*.*07) ELISA]*

**Figure 6.**
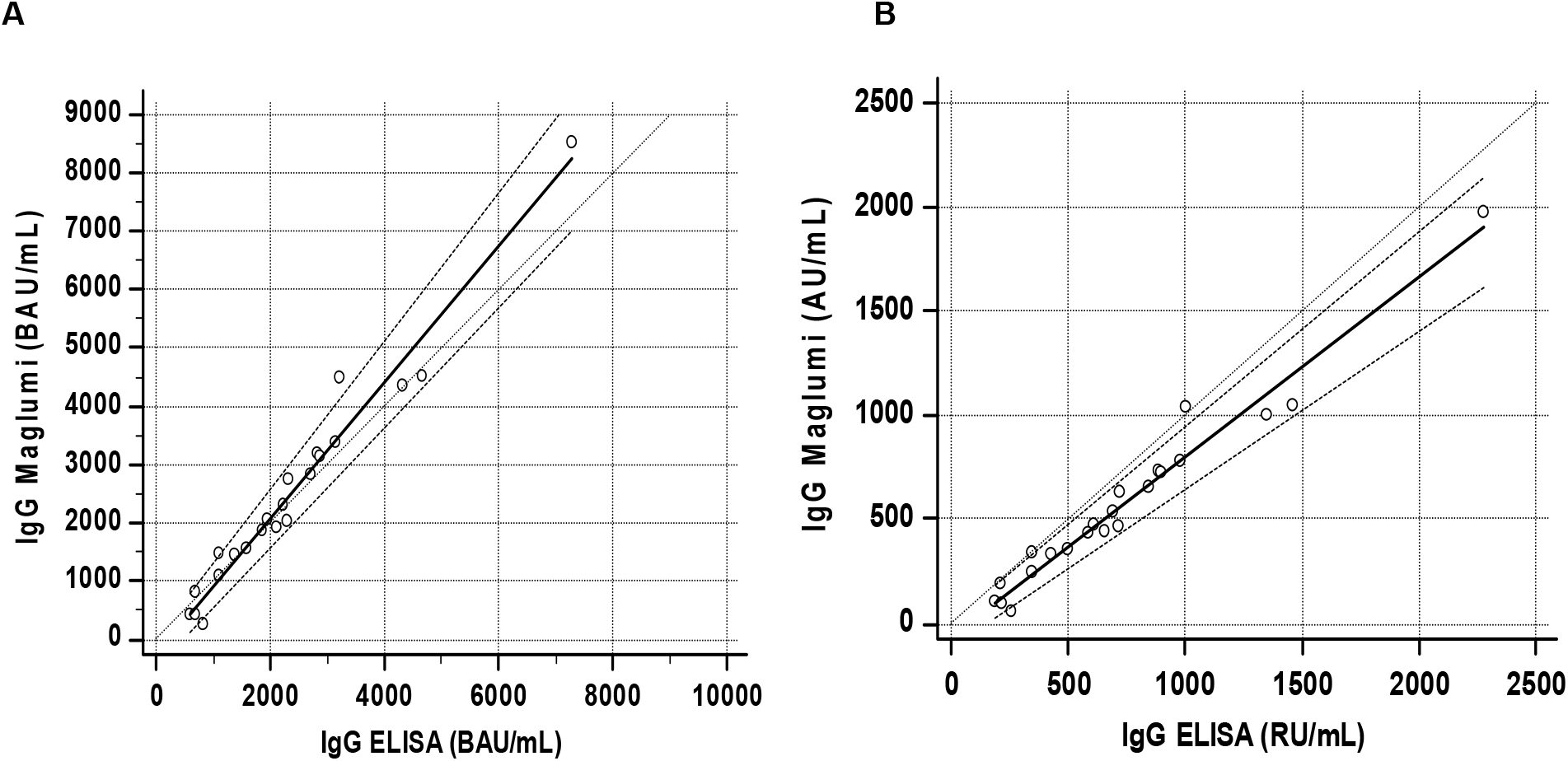
Correlation between ELISA and Maglumi methods in vaccinated subjects 15 days after the second dose. The trend lines represent the Passing-Bablock correlation **A** Concentrations expressed in Arbitrary Units [*Maglumi*= *- 52*.*4(−107*/+*19*.*2)* +*0*.*85 (0*.*74*/*0*.*92)]* **B** Concentrations expressed as BAU [*Maglumi* = *-227*.*9 (−464* / + *69*.*9)* + *1*.*14 (0*.*99* / *1*.*25) ELISA]*

Three months after the second dose the levels of antibodies drastically decrease (table 3). The median of the percentage of the concentrations compared to those found 15 days after the second dose was 21% (10°-90° perc 11-33%) with Maglumi and 24.4% (10°-90° perc 13-33%) with ELISA.

The Passing-Bablock regression between the two methods was: *Maglumi= -16*.*2(−37*.*5*/+*5*.*7)* +*0*.*78 (0*.*65/0*.*98) ELISA* (Fig. 7).

**Figure 7.**
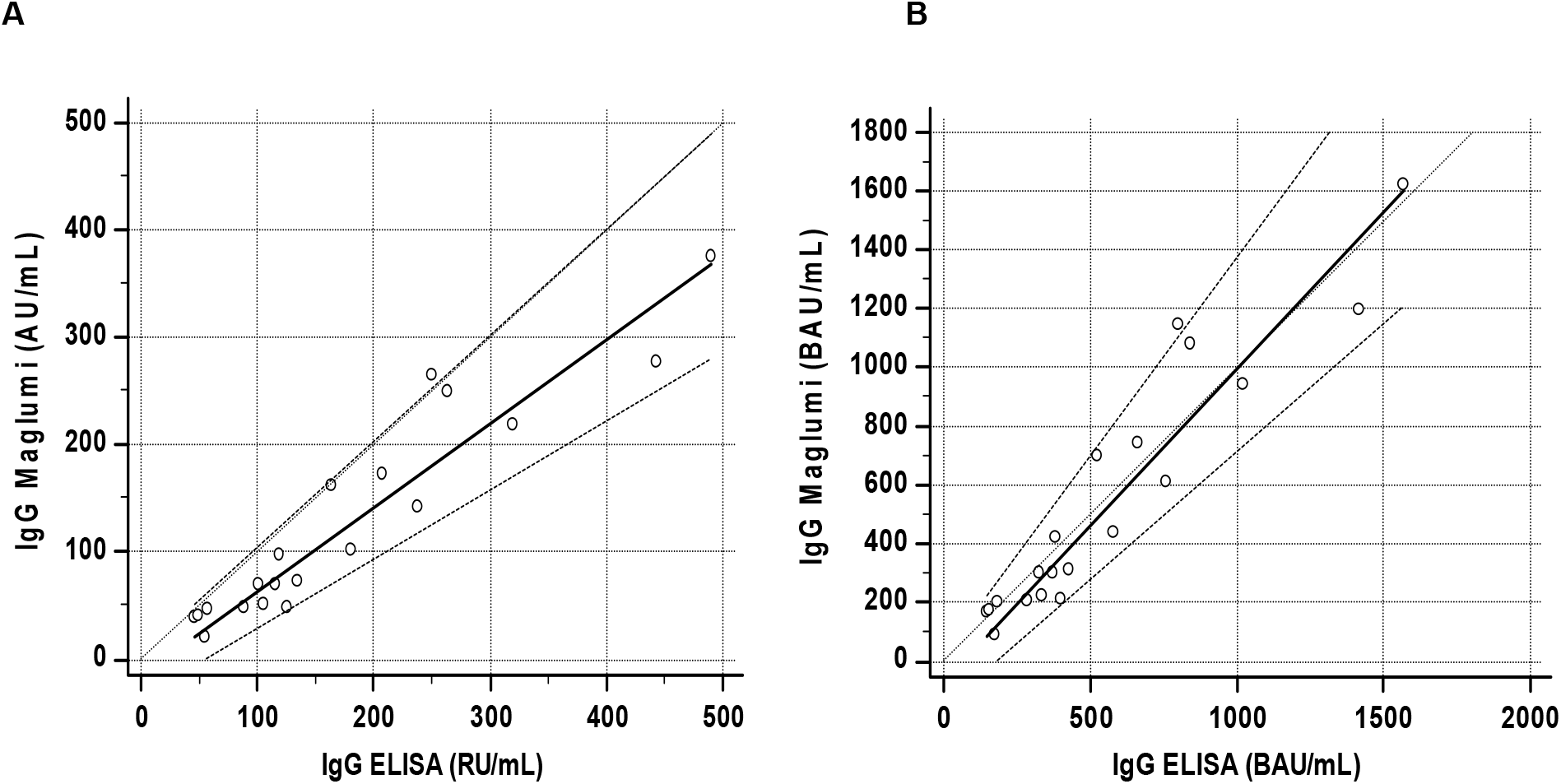
Correlation between ELISA and Maglumi methods in vaccinated subjects 3 months after the second dose. The trend lines represent the Passing-Bablock correlation **A** Concentrations expressed in Arbitrary Units [*Maglumi*= *-16*.*2(−37*.*5*/+*5*.*7)* +*0*.*78 (0*.*65*/*0*.*98) ELISA]* **B** Concentrations expressed as BAU [*Maglumi* = *-72*.*8 (−166*.*8* / + *24)* + *1*.*07 (0*.*88* / *1*.*35) ELISA]*

Correction factors versus the international standard WHO 20/136 were determined for both assays (4.33 for Maglumi and 3.2 for ELISA). The correlations between the two methods after transformation into Binding Arbitrary Units (BAU) resulted in *Maglumi = 2*.*45 (−1*.*6* / + *10*.*6)* +*0*.*39 (0*.*33* / *0*.*46) ELISA* in patients, *Maglumi* = *-5*.*1 (−26*.*8* / + *5*.*0)* +*0*.*82 (0*.*64* / *1*.*07) ELISA* in subjects vaccinated after the first dose and *Maglumi* = *-227*.*9 (−464* / + *69*.*9)* + *1*.*14 (0*.*99* / *1*.*25) ELISA* in subjects vaccinated after the second dose. Three months after the second dose the correlation is *Maglumi* = *-72*.*8 (−166*.*8* / + *24)* + *1*.*07 (0*.*88* / *1*.*35) ELISA*.

## 4. Discussion

In this study, the performance of two assays for the determination of antibodies anti SARS-CoV2 in patients with samples up to 10-12 months from the onset of symptoms were compared. With both methods, antibodies concentrations tend to decline after a few months, but the levels persist relatively high up to nearly a year after symptoms (Tab.2). Our data, especially those obtained with Maglumi, were approximately in accord with a model of the IgG anti-S decay in patients, that established a half-life of 229 days [17]. Positivity rates remains 100% with the Maglumi method, while they drop to 87% with the ELISA method.

In vaccinated subjects, the presence of high concentrations of antibodies is already detectable after the first dose, but after the booster they show a significant increase, about 20 times compared to the first administration and on average 3 times the maximum concentrations reached by the patients after about two months from the onset of symptoms. This result is in agreement with previous findings for the method used [13,18,19]. However, in vaccinated subjects the decrease in antibody concentrations is more rapid, given that after 3 months after the second vaccination they are reduced to less than a quarter. The correlations between the two methods are always acceptable, but while in the patients the results are scattered and the ELISA method has 3-4 times higher levels of Maglumi, in vaccinated subjects the concentrations between the two methods are closer and much better correlated. Moreover, the conversion of the results into BAU units improves the relationship between the two methods. However, only in vaccinated subjects there was no evidence of proportional error after the conversion, while in the patients the difference between the two methods remained significant. The methods measure antibodies directed against the Spike 1 protein, but Maglumi more specifically determines antibodies against the receptor domain. This difference may partly justify the results in the patients. Considering that only Ab anti Spike should be expressed in vaccinated patients, it could be speculated that the greater heterogeneity of antibodies pattern in patients could cause the less close correlation between the two methods found in these subjects. However, the decrease in concentrations a few months after vaccination does not necessarily mean a reduction in protection. It is in fact possible that the protection is not directly proportional to the mere presence of antibodies, given the persistence of T-cell memory after infection [6]. In conclusion, both methods have comparable behaviors, both in patients and in vaccinated subjects. In both cases the antibody concentrations peaked and then decrease. However, in vaccinated subjects the peak reached a much higher levels than in the patients, and the decrease was more rapid. Three months after the second injection they showed concentrations comparable to those of patients after more than 6 months from the onset of the disease. A peculiar finding of the study was the failure of the BAU conversion in the harmonization of different methods only in patients’ specimens. Further studies will be required to clarify the different behavior between patients and vaccinated subjects.

## Data Availability

No data

## Author Contributions

Conceptualization, R.D. and P.C.; sample collection, R.D., I.B. and H.A.; methodology, M.S. I.B. and H.A..; formal analysis, R.D.; writing—original draft preparation, R.D.; writing—review and editing, R.D., M.S., H.A. and P.C.; supervision, P.C. All authors have read and agreed to the published version of the manuscript.

## Institutional Review Board Statement

The study was conducted according to the guidelines of the Declaration of Helsinki.

The Ethical committee for clinical trials, ULSS3 Serenissima, Venice, approved the study (Approval n.149/A CESC).

## Acknowledgments

We acknowledge Medical Systems and Euroimmun for kindly supplying reagents without any influence in study design and data analysis.

